# Python-Streamlit web application to enhance evidence-based medicine education for first year medical students

**DOI:** 10.64898/2026.08.23.26361151

**Authors:** Venkata Patchigolla, Amolak S. Jhand, Hyeok Jun Lee, Laura J. Benjamins

## Abstract

Evidence-based medicine (EBM) concepts are difficult for medical students to grasp. We developed a Python-Streamlit web application providing interactive visualizations to enhance EBM education. Preliminary use with first year medical students demonstrated high engagement and improved conceptual understanding, supporting the feasibility of integrating interactive, web-based tools into EBM curricula.

## Manuscript text

Evidence-based medicine (EBM) demands the integration of the best current research with clinical expertise and patient values to optimize patient outcomes [1]. As a cornerstone of modern clinical practice, it is essential for healthcare professionals to develop proficiency in its application. Effective use of EBM requires not only understanding scientific literature but also accurately interpreting data and statistics from the available literature, an area where proficiency often lags.

Recent online surveys have demonstrated poor statistical understanding among learners despite high self-reported confidence, a phenomenon described as the “illusion of knowledge” [2]. In parallel, a meta-synthesis found that learners prefer structured formats such as lectures, tutorials, and workshops for learning EBM concepts, while the integration of online interactive tools remains in its early stages [3]. To address this gap, the Data Science in Medicine Club at Wayne State University School of Medicine developed a web-based application using the Streamlit library in Python to provide interactive visualizations of core EBM concepts [4]. At our institution, EBM is introduced during the first-year curriculum through a problem-based learning framework consisting of lecture, small-group problem solving, and group discussion. The web-based application was designed to supplement the small-group component by incorporating interactive visualizations to enhance conceptual understanding.

The application (https://dsimctool.streamlit.app/EBM) includes modules covering foundational statistical and epidemiologic concepts, including hypothesis testing, Type I and Type II errors, statistical power, bias, risk and odds ratios, number needed to treat, and measures of risk reduction. Interactive features allow learners to dynamically adjust parameters such as sample size, effect size, and decision thresholds to visualize changes in confidence intervals, sensitivity– specificity tradeoffs, and other relationships that are often difficult to conceptualize. For example, one module demonstrates how increasing sample size narrows confidence intervals, reinforcing statistical intuition through real-time visual feedback [5].

The application was integrated into existing small-group EBM sessions, where students used the tool alongside structured worksheets. These worksheets guided learners through scenarios involving hypothesis testing and error interpretation while interacting with the visualizations. The tool was accessible online, enabling both in-session use and optional independent exploration.

Preliminary evaluation of the application was conducted among a cohort of 300 first-year medical students following a small-group session in which the tool was implemented. Of these, 118 students completed a post-use survey. Engagement was high, with 94% of respondents reporting active use of the tool to complete assigned activities. Survey results are summarized in Table 1. The majority of learners reported that the application enhanced their understanding of EBM concepts and improved their ability to visualize topics they previously found difficult. The tool was also perceived as useful during small-group learning, with high overall satisfaction ratings (66 students very satisfied, 39 somewhat satisfied, 11 neither satisfied nor dissatisfied, 1 somewhat dissatisfied, and 1 very dissatisfied).

**Table 1.** Anonymous post-survey responses following use of the web application following a small group session.

|  | “I feel that the tool enhanced my understanding of EBM topics” | “I feel that interacting with the EBM tool was useful for my learning during the small group session” | “I feel that the tool helped visualize EBM concepts that I felt were difficult or unsure of” |
| --- | --- | --- | --- |
| Strongly agree | 65 | 68 | 59 |
| Somewhat agree | 38 | 34 | 41 |
| Neither agree nor disagree | 11 | 13 | 18 |
| Somewhat disagree | 4 | 3 | 0 |
| Strongly disagree | 0 | 0 | 0 |

However, intention to use the tool outside structured sessions was mixed, with 27 students reporting they planned to use the tool independently, 54 indicating “maybe,” and 37 reporting no intention to do so. These findings suggest that while the application is effective as a guided learning aid, broader independent adoption may require additional refinement. Future iterations will focus on enhancing the modules through the incorporation of clinically relevant scenarios that guide learners through statistical concepts. For example, the sensitivity and specificity module could be expanded to include a clinically grounded scenario, such as evaluating D-dimer as a diagnostic test for pulmonary embolism, to improve real-world applicability and learner engagement.

In summary, this Python-Streamlit based web application provides a practical and scalable approach to enhancing EBM education. By transforming abstract numerical relationships into intuitive visual representations, the application aligns with learner preferences for structured yet engaging educational formats. Additionally, the simplicity and scalability of the Streamlit framework allow for rapid development and dissemination of similar tools across institutions.

Limitations include reliance on self-reported survey data and the absence of objective performance metrics to assess knowledge gains. Future work will focus on longitudinal evaluation of learning outcomes, expansion of visualization modules, and strategies to promote sustained learner engagement outside of guided sessions.

## Data Availability

All data produced in the present study are available upon reasonable request to the authors

https://dsimctool.streamlit.app/EBM

## Declarations

### Ethics approval

This project was conducted as an educational quality improvement/program evaluation activity using anonymous post-session survey responses. No identifiable participant data were collected.

### Consent to participate

Participation in the post-session survey was voluntary and responses were collected anonymously.

### Data availability

The datasets generated and/or analyzed during the current study are available from the corresponding author on reasonable request.

## References

1. Siwek J. Evidence-based medicine: common misconceptions, barriers, and practical solutions. Am Fam Physician. 2018;98(6):343–344.

2. Lakhlifi C, Lejeune FX, Rouault M, Khamassi M, Rohaut B. Illusion of knowledge in statistics among clinicians: evaluating the alignment between objective accuracy and subjective confidence, an online survey. Cogn Res Princ Implic. 2023;8(1). 10.1186/s41235-023-00474-1

3. Chandran VP, Balakrishnan A, Rashid M, Khan S, Devi ES, Kulyadi GP, et al. Teaching and learning strategies of evidence based medicine: a meta-synthesis of learners and instructors perspective. Clin Epidemiol Glob Health. 2023;21:101280. 10.1016/j.cegh.2023.101280

4. Streamlit. A faster way to build and share data apps. Available at: https://streamlit.io/. Accessed 12 May 2026.

5. Patchigolla V, Jhand AS, Lee HJ. DSIMC Tool. Available at: https://dsimctool.streamlit.app/EBM. Accessed 12 May 2026.

